# Demographic Influence on the Effectiveness of England’s SARS-CoV-2 Policies

**DOI:** 10.1101/2023.04.25.23288871

**Authors:** Maharshi Dhada, Jazmin Labra Montes

**Affiliations:** Institute for Manufacturing, Department of Engineering, University of Cambridge, Cambridge, CB3 0FS, U.K.; St John’s College, University of Cambridge, St Johns St, Cambridge CB2 1TP, U.K.

## Abstract

Health data is key for the development of medicinal drugs, treatments, and policy-planning to control the spread of infectious diseases. However, the collection, curation, and interpretation of health data is often biased. This paper discusses England-wide impact of public policies to control the spread of SARS-CoV-2 (Covid-19) infections, based on the recorded per-capita infection cases between July 2020 to January 2023. The analysis presented herewith highlights the disparities across the upper local tier authorities, in the number of Covid-19 cases recorded in response to the policies. This paper further presents the correlation between the Covid-19 cases count and demographic factors, thus highlighting the key factors determining the effectiveness of the NHS policies, and therefore the need for incorporating demographic imbalance in the policy planning process. It is concluded that the upper local tier authorities comprise of three clusters of low, mid, and high prevalence of Covid-19 infections. Where the regions with high prevalence of Covid-19 cases are also the ones with higher proportions of Black/ Mixed racial groups, amongst a mid-range and low internal Migrations.

## Introduction

Health data refers to physical or mental information about an individual collected in a digital or physical form^1^. Health data often contains information related to health status including future, past and current treatments, causes of deaths, among several other socio-economic parameters^2,3^.

Health data is key for the development of medicinal drugs, treatments, and policy-planning to control the spread of infectious diseases. Clinical treatments^4,5^, facilitating the communication between the health care service sectors^6,7^, and developing the cure of diseases^8,9^ all rely on health data. Additionally, and as recent events have shown, collecting, and understanding data is also crucial for public decision-making^10^, for instance during lockdowns when to close facilities^11^ including schools, universities, gyms, as well as to take measures to prevent and contain novel diseases^12^. Health data is thus a constant factor for the whole health care sector: from establishing bridges between different services, their use for scientific research to public interest, the collection, curation, interpretation, and extrapolation of health data has modernized the medical services.

Nonetheless, the collection, curation, and interpretation of health data is often biased. The bias is specifically disadvantageous to the minority sectors, making them invisible in policy-planning for the broader population. A long-standing problem and as recognised by the World Health Organization is that health data has historically been focused on “privileged people”^13^. Since health data is used as input for the taking of decision and their impact on the population^14^, the development of drugs, treatments and prevention of side-effects^15^, the lack of data from minorities makes vulnerable sectors invisible and reinforce the gap between privileged people and population from struggling backgrounds^16^.

There are two types of local authorities in England: county councils and district councils. Upper-tier local authorities (UTLAs) are county councils, which are responsible for services such as education, social services, highways and transport, and waste management, among others. One of the responsibilities of the UTLAs is to uphold public health, where they are responsible for providing health services, such as immunisation programs and health education. During the Covid-19 played an especially critical role in developing and implementing policies to control the spread of infections. They were particularly involved in developing and implementing local outbreak control plans, coordinating local responses, providing public health advice and support, and monitoring compliance with regulations.

Covid-19 polivies in England comprised of lockdowns, social distancing measures, mask mandates, and vaccination campaigns.

In March 2020, the UK government introduced a national lockdown in response to the COVID-19 pandemic. This lockdown required people to stay at home except for essential purposes such as buying food and medicine or exercising. This policy was successful in reducing the spread of the virus, and in June 2020, the government began to ease restrictions. However, the number of cases began to rise again, and in November 2020, a second national lockdown was introduced.

In December 2020, the government announced that it had approved the Pfizer-BioNTech COVID-19 vaccine for use in the UK. The vaccination campaign began shortly afterward, with priority given to the elderly, healthcare workers, and people with underlying health conditions. By April 2021, over 32 million people in the UK had received their first dose of the vaccine.

In early 2021, the government introduced a tiered system of restrictions based on the prevalence of the virus in different areas. This system allowed for some businesses to reopen and for people to meet in outdoor spaces with limited numbers. However, in January 2021, a third national lockdown was introduced, with schools closed and people once again being asked to stay at home except for essential reasons.

Overall, the role of upper-tier local authorities in COVID-19 policies has been to provide local leadership and coordination in the response to the Covid-19 pandemic, working closely with national government and other local organizations such as the National Health Service (NHS) to control the spread of the virus and protect public health. Figure 1 shows the UTLAs and 7 NHS regions across England.

**Figure 1.**
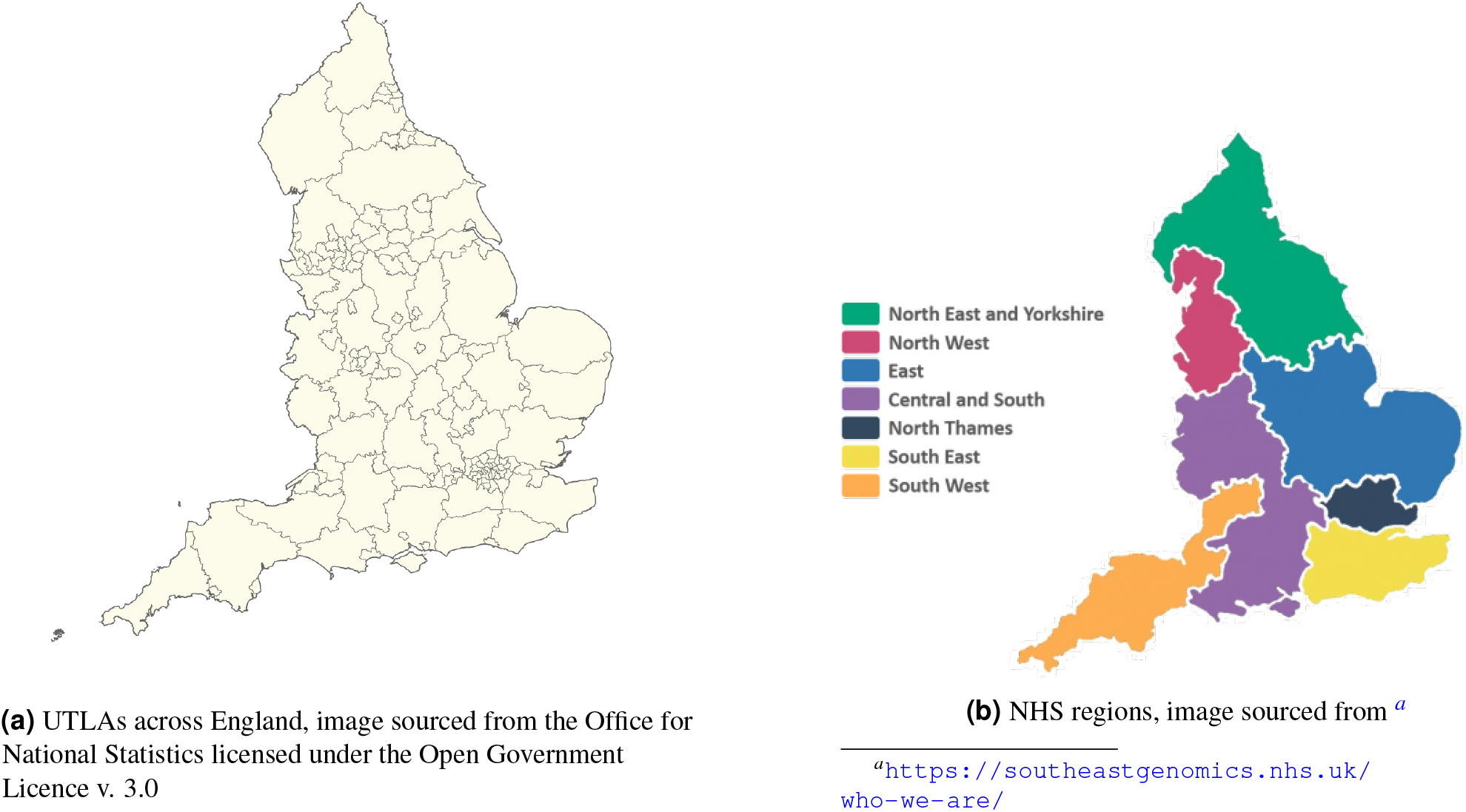
Maps showing various UTLA and NHS regions

The COVID-19 policies implemented in England have had significant impacts on people’s lives, both positive and negative. On the one hand, lockdowns and other restrictions have been successful in reducing the spread of the virus and saving lives. The vaccination campaign has also been successful in reducing the number of hospitalizations and deaths.

On the other hand, the policies have had a severe impact on the economy, with many businesses forced to close or reduce their operations. The lockdowns have also had negative impacts on mental health, with many people experiencing isolation, loneliness, and depression. The closure of schools has also had a significant impact on children’s education and development. The policies have also highlighted existing inequalities in society, with some groups, such as low-income families and ethnic minorities, being disproportionately affected by the virus and the policies aimed at controlling it. The pandemic has also highlighted the importance of a strong social safety net and adequate support for vulnerable groups.

To that end, this paper discusses England-wide impact of governmental policies to control the spread of Covid-19 infections, based on the recorded per-capita infection cases between July 2020 to January 2023 obtained from the official NHS website ^1^. The analysis presented herewith highlights the disparities across the upper local tier authorities of the number of per-capita cases recorded in response to the NHS policies. The disparities are highlighted by using Self-Organising Maps (SOMs) to cluster the timeseries. This paper further presents the correlation between the Covid-19 cases count and demographic factors, thus uncovering the key factors determining the effect of the NHS policies and the need for incorporating demographic imbalance in the policy planning process. This is done by using clusters obtained via using the clusters identified using SOM as targets and demographic data as features, to train a Random Forest classifier. Permutation and Shapley importance scores were used to evaluate the determining features. The most important features identified through this process where further analysed for each of the clusters. The demographic data is obtained from the UK’s National Statistics Website ^2^.

Results following the analysis demonstrate that the upper local tier authorities comprise of three clusters of low, mid, and high prevalence of Covid-19 infections. Moreover, the regions with higher prevalence of Covid-19 cases are also the ones with higher proportions of Black/ Mixed racial groups, amongst a mid-range and low internal Migrations.

## Results and Methods

### Disparities in Covid-19 Infections across England

Figure 2 presents the Covid-19 cases recorded in the UK, across its upper local tier authorities. In Figure 2a, the shaded portion represents the first standard deviation (along y-axis) of Covid-19 infections per-100,000 population, and the corresponding mean. It is clear that there is a significant variation across the UTLAs, in terms of number of Covid-19 infections. to further highlight this variation, Self Organizing Map (SOM) was used for clustering the timeseries into three difference clusters representing high, moderate, and low number of Covid-19 infections explained in the following subsections. This variation in the observed number of cases is also evident for various NHS regions, shown in Figure 2b.

**Figure 2.**
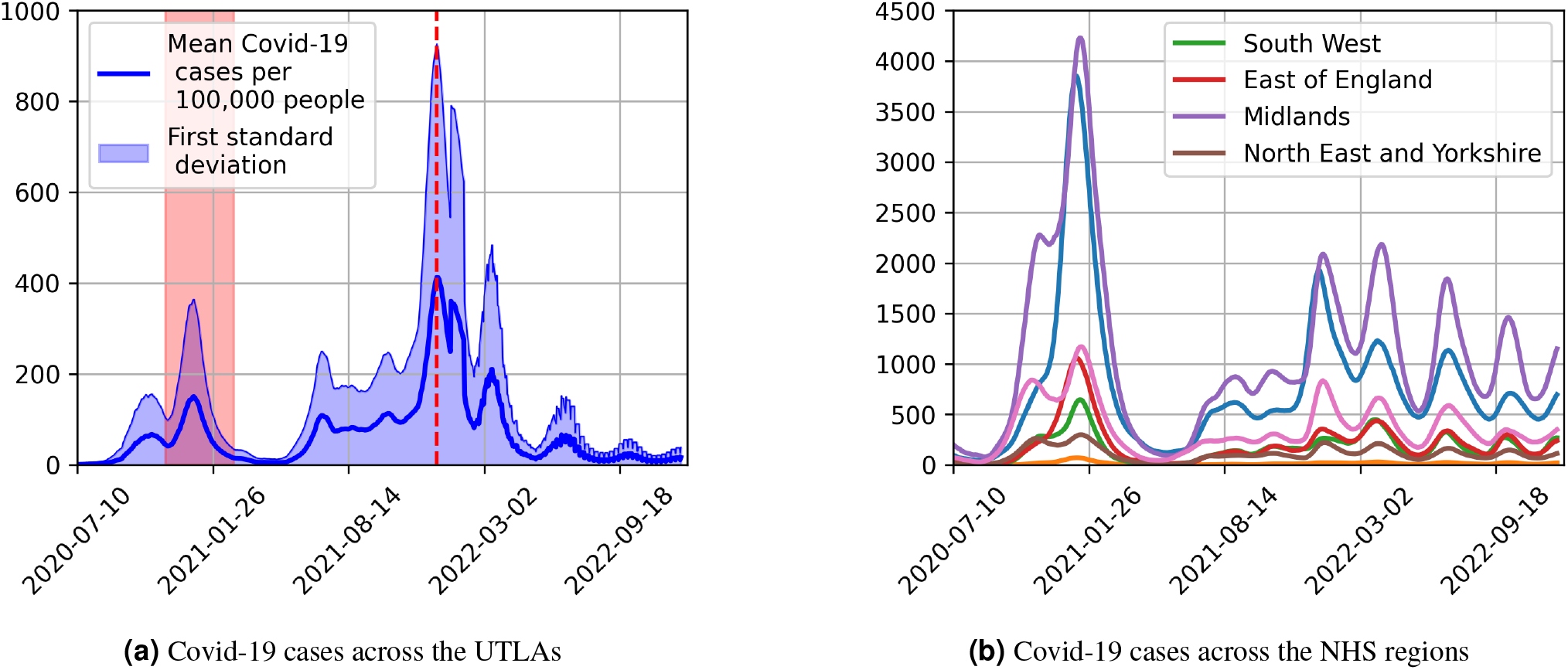
Timeseries showing the Covid-19 cases recorded across the UK

#### Self Organizing Maps for Timeseries Clustering

Self-Organizing Maps (SOMs) are a type of artificial neural network that can be used for unsupervised learning and clustering. They are particularly useful for analyzing high-dimensional data, including time-series data, by projecting it onto a lower-dimensional grid. In particular, SOMs can be a powerful tool for time-series clustering that can be used to identify patterns and relationships in complex time-series data. By projecting the data onto a two-dimensional grid, SOMs can visualize and explore the underlying structure of the data and to identify groups of similar time-series data based on their patterns and characteristics. To apply SOMs to time-series data, the input data is first transformed into a sequence of feature vectors, with each vector representing a different time step in the series. The SOM is then trained using these feature vectors to create a two-dimensional grid of neurons that represents the patterns in the data. During training, the SOM adjusts the weights of each neuron to represent different regions of the input space, with neurons that are close to each other in the grid representing similar input patterns.

Here, for time-series clustering using SOMs, a modified distance measure called Dynamic Time Warping (DTW) distance that takes into account the temporal relationships between the feature vectors is used. DTW measures the distance between two time-series data by aligning their sequences in time and calculating the minimum distance between corresponding points.

The weight update equation for SOMs is modified to incorporate the temporal relationships between the feature vectors as follows:

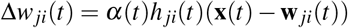

where Δ*w*_*ji*_(*t*) is the change in weight for neuron *j* at time *t, α*(*t*) is the learning rate at time *t*, **x**(*t*) is the feature vector at time *t*, and **w** _*ji*_(*t*) is the weight vector for neuron *j* at time *t*. The function *h* _*ji*_(*t*) is the neighborhood function that determines the degree to which neighboring neurons influence the weight update of neuron *j* at time *t*.

In this case, 50,000 iteration was deemed sufficient as the algorithm converged after this.

Figures 4 and 5 present the cluster counts and timeseries in each of the clusters. It should be noted that although majority of the cities belong to lower recorded Covid-19 infections, there are UTLAs where significantly higher instaces of Covid-19 infections are observed.

**Figure 4.**
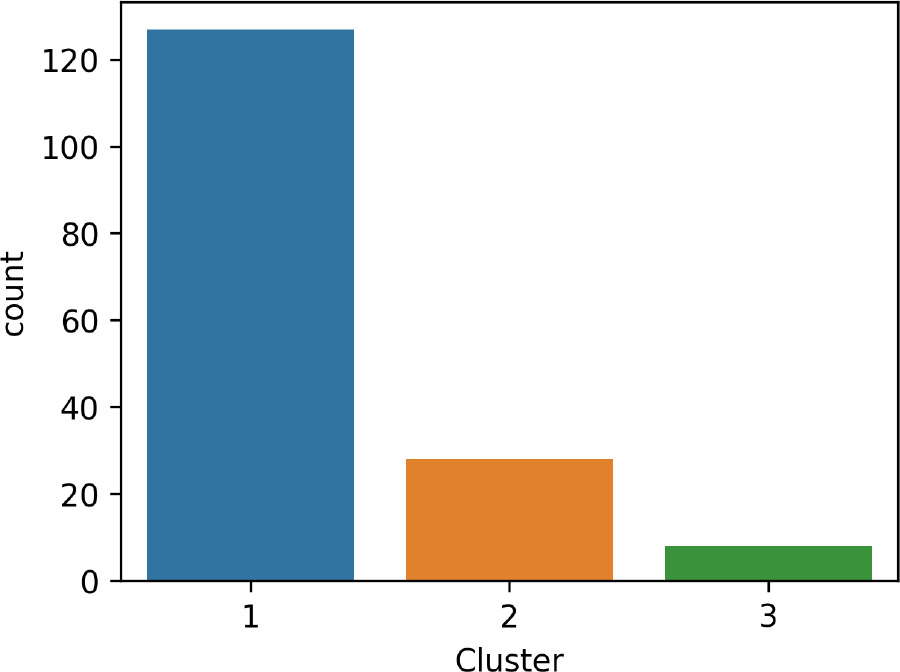
Number of UTLAs per cluster

**Figure 5.**
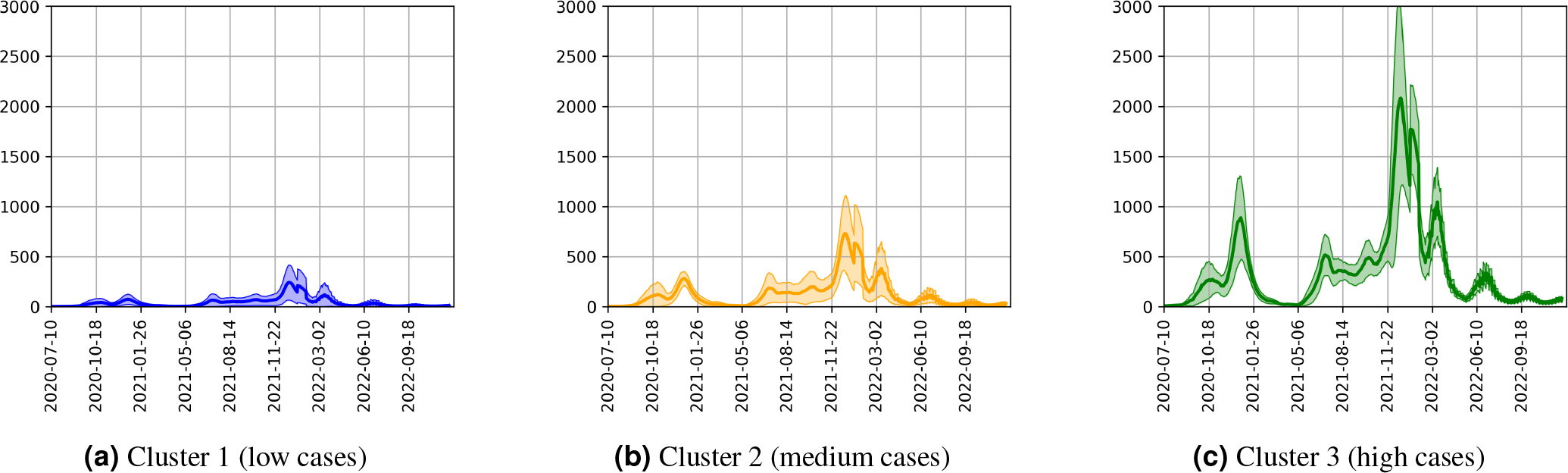
Number of cases within each cluster

### Feature Analysis

Using the clusters of UTLAs identified above, corresponding to low, moderate, and high Covid-19 infections, a Random Forest Classifier was trained using the above clusters as labels and demographic data as features.

More specifically, the following demographic features, corresponding to a UTLA, were used:

1. **Population Age**:
2. **Ethnicity**
3. **Gender Diversity**
4. **Migration**
5. **Population Density**

#### Random Forest Classifier

Random Forest (RF) is an ensemble learning method that is used for classification, regression, and other tasks. In RF, multiple decision trees are trained on different subsets of the data and their results are combined to make predictions. Each tree in the forest is constructed using a random subset of the features and a bootstrapped sample of the data.

The algorithm works as follows:

1. A random subset of features is selected.
2. A decision tree is trained on the bootstrapped sample of the data using the selected features.
3. Steps 1 and 2 are repeated multiple times to create a forest of decision trees.
4. To make a prediction, the input data is passed through each tree in the forest, and the results are aggregated to produce a final prediction.
5. The final prediction is determined based on the majority vote or average of the results from all the trees in the forest. RF reduces overfitting by introducing randomness and variability in the tree building process.

The RF algorithm can be expressed mathematically as follows:

Let *T* be the number of trees in the forest, and *h*_*t*_(*x*) be the prediction of tree *t* for input *x*. The prediction for the RF ensemble is given by:

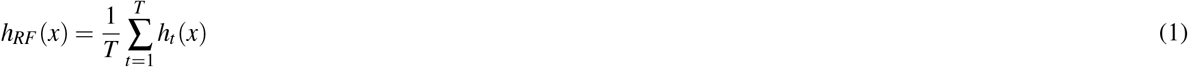

Where *h*_*RF*_ (*x*) is the prediction for input *x* using the RF ensemble.

RF classifiers are widely used in machine learning due to their high accuracy and ability to handle large datasets with many features. They have applications in fields such as finance, medicine, and natural language processing.

For the current analysis, an RF comprising 2000 decision trees using four features at once with aggregate bagging of the dataset was used. An example of a decision tree used in the current analysis is shown in Figure 6

**Figure 6.**
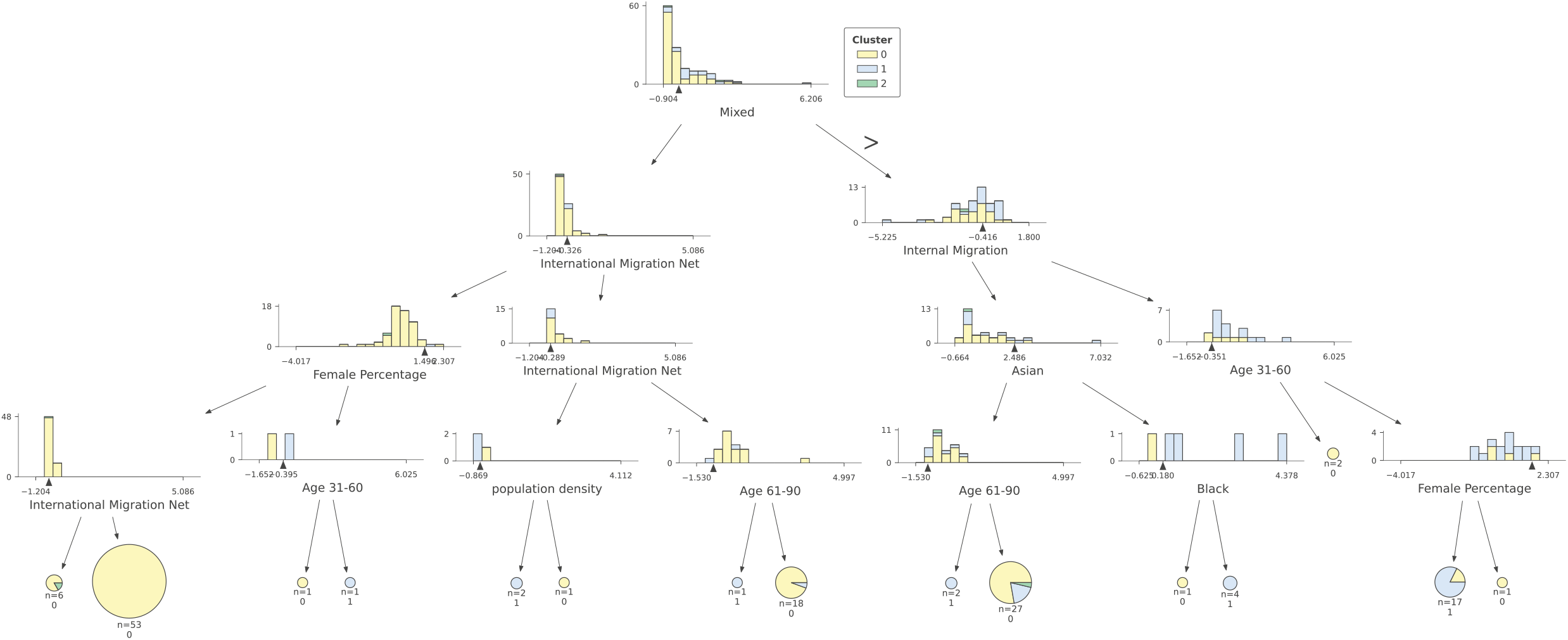
An example decision tree

**Figure 7.**
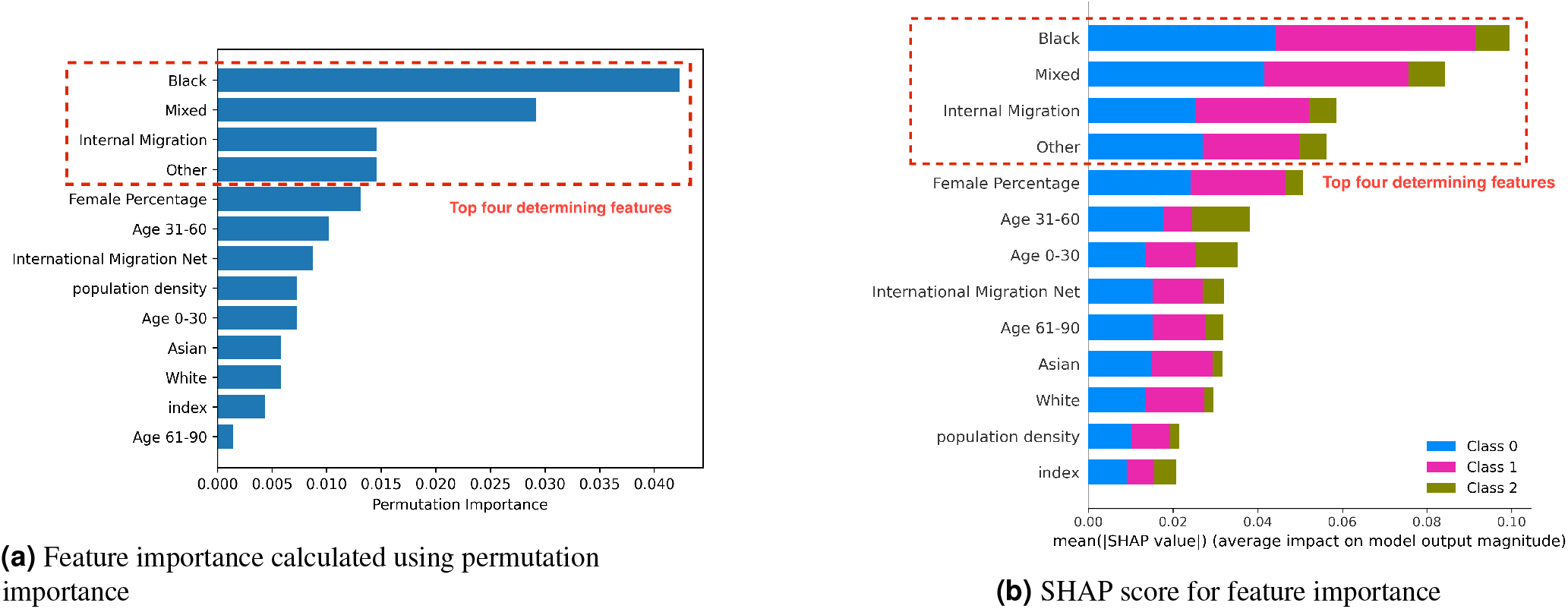
Feature importance determined using the permutation importance and SHAP scores

The following metrics were used to quantify the feature importance, given the above identified timeseries clusters as targets. In other words, these numbers show the importance of each feature to classify the corresponding UTLA into the possible classes, and in turn the effectiveness of NHS policy in that area.

#### Permutation Feature Importance

Permutation feature importance is a method for feature selection in machine learning models. It measures the importance of a feature by randomly permuting its values and measuring the effect on the model performance. The permutation feature importance for categorical variables can be computed as follows:

Let *X* be the categorical feature of interest and *y* be the target variable. The model is trained using *X* and *y*, and its performance is evaluated using a performance metric *M*. The permutation feature importance for *X* is then defined as:

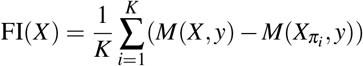

where FI(*X*) is the feature importance score for *X, K* is the number of permutations, 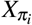 is the feature *X* with its values randomly permuted in the *i*-th permutation, and *M*(*X, y*) and 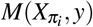 are the model performance scores for the original feature and the permuted feature, respectively.

The permutation feature importance score for *X* measures the effect of permuting the values of *X* on the model performance. A high importance score indicates that the feature is important for the model, and a low score suggests that the feature is not relevant.

#### Shapely (SHAP) Value for Feature Selection

Shapley value is a concept from cooperative game theory that has been applied to feature selection in machine learning. It measures the contribution of each feature to the model performance by computing the marginal contribution of a feature to the overall performance. The Shapley value for a categorical feature is computed as follows:

Let *X* be the categorical feature of interest and *y* be the target variable. The Shapley value for *X* is defined as:

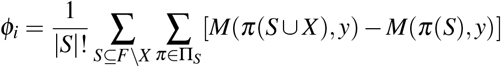

where *ϕ*_*i*_ is the Shapley value for feature *X*, |*S*| is the size of the subset *S* of features excluding *X*, Π_*S*_ is the set of all possible orderings of the features in *S*, and *M*(*π*(*S* ⋃ *X*), *y*) and *M*(*π*(*S*), *y*) are the model performance scores for the ordered feature set including and excluding *X*, respectively.

The Shapley value measures the contribution of a feature to the model performance by averaging the marginal contributions of the feature to all possible subsets of features in the model. A high Shapley value indicates that the feature is important for the model, and a low value suggests that the feature is not relevant.

#### Analysing the Features with Highest Scores

**Figure 8.**
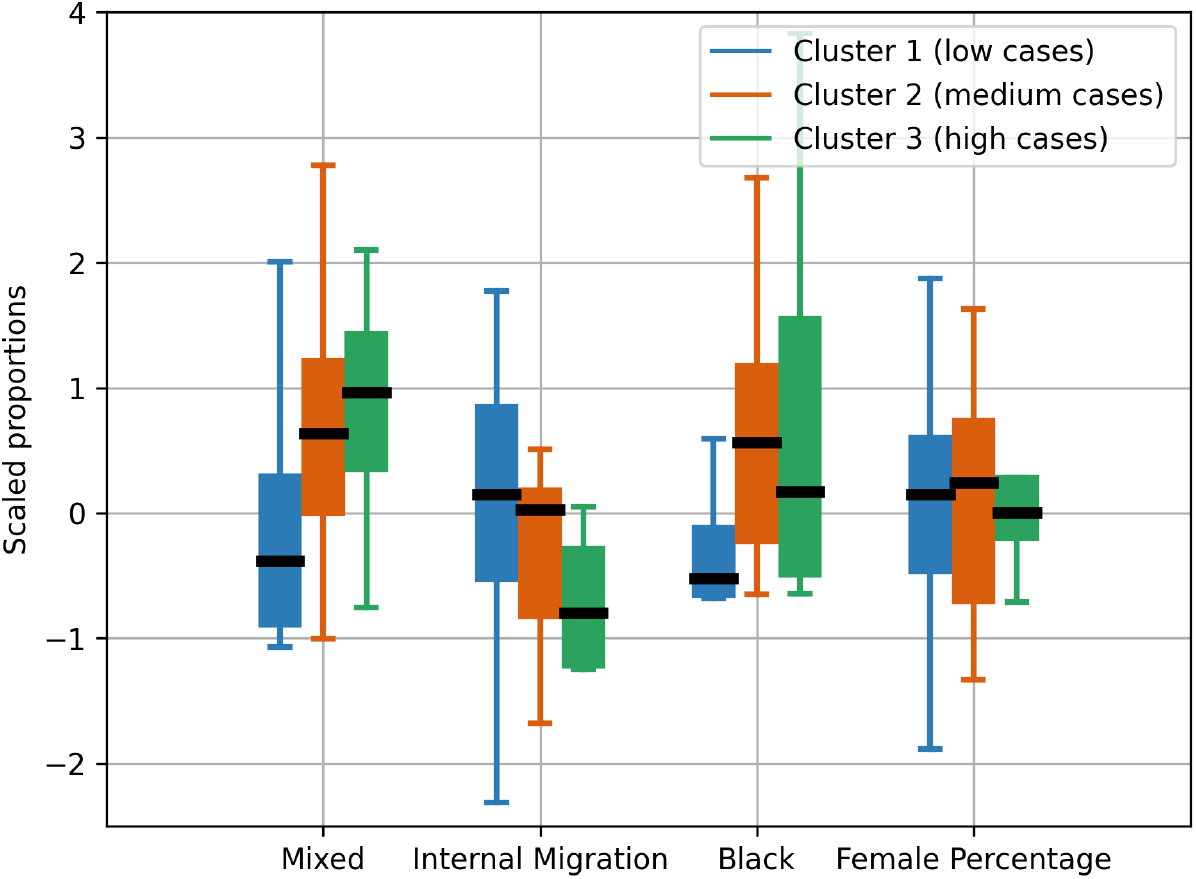
Comparison of the top features responsible for determining the policy effectiveness

## Discussion

The results presented above show the NHS policies do not apply evenly to the UK population. This is evident by clusters of timeseries observed in the recorded Covid-19 cases across various upper tier local authorities considered in this study.

The analysis presented herewith demonstrates that policy making should be align with theories, that generic policies are not the best way to pursue improvements and changes in the society. By taking into consideration the unique characteristics of its population, health agencies and governments can induce more case effective policies and strategies with the aim to stop the spreading of diseases.

## Methods

## Data Availability

All data produced in the present work are contained in the manuscript

## Additional information

The author(s) declare no competing interests.

## Footnotes

1 https://coronavirus.data.gov.uk/details/download

2 https://www.ons.gov.uk/peoplepopulationandcommunity/populationandmigration/populationestimates/datasets

